# From Dental to Medical Imaging: Translational 8 μm Pixel Size, Low-dose and Ultra-High-Definition X-ray Detector for Microfocus Clinical Applications

**DOI:** 10.64898/2025.12.01.25341101

**Authors:** D Uzbelger Feldman, E Simons, R Turchetta, A Bofill-Petit, RJ Raible

## Abstract

**Background:** Accurate radiology disease detection relies on high-resolution, low-dose imaging, yet current systems frequently fail to identify small lesions early enough to alter prognosis while minimizing radiation exposure. This occurs because existing detector architectures cannot maintain high quantum efficiency at small pixel sizes without increasing radiation output. Radiography (70–100 µm), mammography (50–100 µm), CT (250–500 µm), and cone-beam CT (80 µm) detectors are constrained by pixel size, resulting in limited modulation transfer function (MTF), noise power spectrum (NPS), and relative detective quantum efficiency (rDQE). These limitations contribute to 56% of diagnostic errors and require higher milliampere (mA) settings. The objective of this study was to compare a novel CMOS size-2 intraoral X-ray detector prototype with DR, mammography, CT, and CBCT detectors in terms of dose efficiency and spatial resolution (lp/mm), and to evaluate the feasibility of ultra-high-resolution, low-dose X-ray imaging for medical radiology applications.

**Methods:** We developed and evaluated a complementary metal–oxide semiconductor (CMOS) size-2 intraoral dental and small-field detector prototype comprising a novel back-illuminated (BI) pixel architecture and microlenses (M) to reduce pixel size to 8 µm, enabling low-dose acquisition. MTF, NPS, and rDQE were benchmarked against published data for radiography, mammography, CT, and CBCT in terms of dose efficiency and spatial resolution in line pairs per millimeter (lp/mm). A microfocus X-ray source was used at 70 kVp, 0.3 mA, 20 µm focal spot, 5 cm source-to-detector distance, and 0.25 s exposure time.

**Results:** The delivered dose rate during prototype acquisitions was 83.6 mGy/s at a 5 cm source-to-detector distance, corresponding to an air kerma of 20.9 mGy per image for a 0.25 s exposure (0.075 mAs). Inverse-square scaling indicates that incident air kerma would decrease at typical clinical distances once exposure settings are adjusted to maintain signal and image quality. These measurements therefore demonstrated that, at 0.3 mA and a short geometry, the 8 µm BI-M design improved image resolution to 30 lp/mm, exceeding the 5–10 lp/mm limits of existing modalities, while operating at a radiation output several-fold lower in mAs than current systems, providing dose-efficiency headroom for future clinical configurations. Additional system-level analysis demonstrated that microlenses further improved the measured system MTF across clinically relevant spatial frequencies under scintillator-coupled, low-dose imaging conditions.

**Conclusion:** Although originally developed for intraoral imaging, the novel 8 µm BI prototype detector’s pixel architecture, combined with microlenses and a microfocus X-ray source, enabled higher dose efficiency and spatial resolution compared with radiography, mammography, CT, and CBCT detectors. It surpassed MTF, NPS, and rDQE performance at a lower dose, highlighting its potential for early-stage disease detection with reduced radiation burden in microfocus clinical applications. The 8 µm BI-M pixel architecture broke the traditional dose–resolution trade-off by preserving quantum efficiency at small pixel size and enabling microfocus-based clinical imaging at low mA. These technological improvements are particularly relevant for cancer detection, pediatric imaging, and thin-anatomy applications.

## Introduction

Radiation exposure is becoming the “new tobacco,” a growing public health concern driven by cumulative medical imaging dose. Every year, more than 4.2 billion diagnostic radiology examinations are performed worldwide. There were an estimated 691 million radiologic, computed tomography (CT), dental, and nuclear medicine studies performed in the United States (US) in 2018.^1^ The US also accounted for 74 million CT procedures (18% of the world’s estimated total), 275 million conventional radiology procedures (11% of the world’s total), 8.1 million interventional radiologic procedures (34% of the world’s total), 320 million dental radiography procedures (29% of the world’s total), and 13.5 million nuclear medicine procedures (34% of the world’s total).^2^ Public health research estimates that CT imaging in the US alone may account for > 100,000 future cancers each year.^3^ Epidemiological studies in individuals exposed to medical CT scans suggested that cancer risk may increase even at doses comprising up to five CT scans. One-third of children who have undergone CT scans have at least three scans. Children and teens exposed to radiation during CT scans are 24% more likely to develop cancer, according to a large (680,000 patients) long-term study.^2^ Overutilization also exposes patients to unnecessary radiation, and a 6–10% retake rate increases the average population dose resulting from medical imaging.^4^ In clinical digital radiography (DR), exposures are delivered as short pulses; representative peer-reviewed settings are ∼120 kV at 8–15 mA for ∼18 ms per image (≈0.14–0.27 mAs). In comparison, mammography operates at 100 mA for large focus (100 to 200 mAs), while CT is typically performed at 100–300 mA (≈15–35 mGy; 1.8–3.2 mSv effective dose), CBCT protocols utilize 5–20 mA and yield doses of ∼0.1–0.3 mSv for dental/maxillofacial fields or several cGy for large-FOV IGRT scans, while dental intraoral routines use an average of 7 mA.^5 6 7 8 9 10 11 12 13 2 14 15^ Although ionizing radiation must be controlled in such a way that the doses delivered are as low as reasonably achievable (ALARA), balancing patient benefit with risk,^16^ research studies show that dental X-rays have been associated with an increased risk of brain and salivary gland tumors, thyroid cancer, and low birth weight.^17 18 19 20 21 22 23 24 25 26 27^ In addition, 10% of dental practices and 11.3% of dental schools repeat an X-ray, needlessly overexposing the patients.^28^ In this regard, existing medical and dental X-ray detectors use large pixel sizes to increase the number of photons captured in the photosensitive areas. An advantage of increasing the pixel size is radiation dose reduction, for pulsed and low mA X-ray imaging applications like fluoroscopy. Thanks to their large pixel size, more photons converted by scintillators or the photon-counting pixel can be captured by the detector. Unfortunately, an increase in pixel size proportionally decreases image resolution and disease detection.^29 30^ Research has suggested that as many as 20%–50% of high-tech imaging procedures fail to provide information that improves patient welfare.^2^ Most diagnostic failed detection happens with breast cancer scans, abdominal and pelvic scans, and fractures, and 56% of errors are subtle. Failure to identify fractures may account for 41%-80% of diagnostic errors in the Emergency Department.^2 4^These types of claims account for about 40%-54% of radiology-related medical malpractice cases. Mammography is mentioned in 43% of cases, and X-rays and CT in 19% and 15%, respectively.^31^ General radiography and mammography detectors with 70–100 µm pixel pitches (some as small as 50 µm in newer systems) are limited to ∼5–10 lp/mm. Clinical CT scanners typically use in-plane detector elements of 250–500 µm, constraining resolution to ∼1–2 lp/mm. Cone-beam CT (CBCT) detectors typically have pixel sizes of 80 μm with a 6.25 lp/mm limiting resolution. At these scales, microcalcifications, micro metastases, subtle fractures, or early tumor margins fall below discernibility, contributing to failed detection.^1 2 3 4^ In dentistry, cracked tooth symptoms have become one of the major causes of tooth loss in adults. Diagnosing vertical root fractures (VRF) was among the most challenging tasks for 2D intraoral X-ray radiographs. Compared with intraoral dental radiography, a clinical study of 42 suspected cases in 47 patients indicated that tooth fracture, especially VRF detected by 3D cone beam computed tomography (CBCT), showed significantly higher diagnostic accuracy.^32^ However, the voxel size of the CBCT was reported to be around 75–400 μm, which made the diagnosis of the finer micro-crack impossible. Intraoral X-ray imaging sensors have been optimized for caries and periodontal disease diagnosis to an average of 19 μm pixel size. Despite these efforts, decay can only be detected after attacking 30-40% of the enamel due to the 8-um size of the tooth’s enamel rods.^33^ Also, the smaller pixel size doesn’t allow radiation dose reduction for fluoroscopy modalities in dentistry at the standard 7 mA x-ray tube setting.^34 35^ These shortcomings reflect fundamental design limitations of conventional front-illuminated complementary metal-oxide semiconductors (CMOS) based x-ray detectors, where pixel size and architecture directly constrain spatial resolution and, in turn, diagnostic accuracy. In a front-illuminated design, the photosensitive area lies beneath multiple metal interconnect and transistor layers, reducing the fill factor and quantum efficiency. This configuration is often compared to the human eye, where the light-sensitive retina is located behind vasculature and supporting tissue, so incident photons must traverse additional layers before detection. Thus, both pixel–size–driven mA dose burdens and resolution limits are central to the problem of radiographic failed detection. These dose-related and resolution-related limitations form two sides of the same constraint: current detectors reduce dose by increasing pixel size, but larger pixels inherently degrade resolution. The objective of this study was to compare a novel CMOS size 2 intraoral X-ray detector prototype (Real Time Imaging Technologies, LLC, Charlotte, NC) to DR, mammography, CT, and CBCT medical detectors in terms of dose and resolution in lp/mm and to evaluate the feasibility of ultra-high resolution, low-dose X-ray imaging in medical radiology.^36^

## Methods

The prototype detector consisted of a CMOS sensor with 8 µm pixels, fabricated on a novel back-illuminated (BI) pixel architecture with microlenses (M). BI CMOS sensors invert the front-illuminated architecture, placing the photosensitive layer directly at the entrance window, eliminating obstruction and dramatically improving quantum efficiency. It is like having the retina in the front region of the eye. Combined with M arrays for light guiding into the pixel, BI designs maximize photon capture, enabling smaller pixel sizes while maintaining high signal-to-noise performance (Figure 1).^36 37 38 39^ The overall design emphasized both small pixel pitch and efficient light collection, enabling extension of usable resolution well beyond that of existing detectors. A high-resolution, 0.5 mm thick Cerium-doped Lutetium Yttrium Silicate (LYSO (Ce)) scintillator (Epic Crystal Co. Ltd., Jiangsu, China) was mechanically coupled to the sensor to convert incident X-ray photons into visible light. A microfocus X-ray source (uXR-130 M108, X-ray Vacuum Technology, Co., Ltd., Jiangsu, China) was employed as the radiation source with no need for magnification techniques. Operating parameters were set to 71.7 kVp, 0.3 mA beam current, 2.5 mm Al HVL filtration, 0.5 seconds exposure time with a 20 µm focal spot to minimize geometric blur and penumbra relative to the detector resolution.^40^ A resolution test pattern (Pacific Northwest X-ray Inc., Gresham, OR) was placed next to the detector at a distance of 5 cm from the X-ray source. Image noise reduction was achieved via a customized application comprising mathematical algorithms and convolutional neural network denoising (Real Time Imaging Technologies, Charlotte, NC). Resolution was confirmed by means of ImageJ (U.S. National Institutes of Health, Bethesda, Maryland, USA) plotting. The modulation transfer function (MTF) was measured using a slanted-edge technique on a tungsten bar. The image was then cropped into a 512×512 pixel region of interest (ROI) in accordance with the IEC 62220-1:2015 methodology and analyzed using ImageJ. Edge spread functions were oversampled and windowed to derive line spread functions and the Fourier MTF. The noise power spectrum (NPS) was calculated from a full-frame (3000×3864 pixels) ROI. To reduce readout variability, flat-field acquisitions consisted of a 32-frame sum. The relative detective quantum efficiency (rDQE) was computed from the slanted-edge 512×512-pixel ROI in ImageJ. DQE is reported on a relative scale; absolute DQE was not estimated. Typical detector characteristics for conventional systems are DR 70–100 µm (Nyquist ≈ 7.14 lp/mm), mammography 50-100 um (Nyquist ≈ 5-10 lp/mm), CT 250–500 µm (Nyquist ≈ 1.25 lp/mm), and CBCT 75–80 µm (Nyquist ≈ 6.67 lp/mm). Published measurements from these modalities are cited in the following texts as benchmarks. For flat-panel DR, Alikunju et al. (2023) report pMTF and DQE curves that depend on CsI thickness and dose (see Fig. 10 for pMTF, Fig. 11 for NNPS, Fig. 12 for DQE). For mammography, García-Mollá et al. (2011) reported MTF, DQE, and NPS curves and dose (Figs. 1-5) comparing different 100 μm pixel size flat panel detectors.^41^ For multi-slice CT, Monnin et al. (2020) provide TTF, NPS, and system DQE across reconstruction kernels and dose (Fig. 2, Fig. 4, Fig. 7). For CBCT, Jaffray & Siewerdsen (2000) report DQE(0) as a function of detector configuration and exposure (Fig. 12), and Steiding et al. (2014) provide representative MTF and NPS measurements for dental CBCT systems (Fig. 7 and Fig. 5). Specific numeric comparisons (e.g., DQE(0) for DR or CBCT) are cited directly to those figures in the main text. ^5 9 12 41 42^ System MTF was measured using a tungsten bar target tilted by 10°, imaged with the LYSO(Ce) scintillator at 70 kVp, 0.3–0.5 mA, 0.25 s exposure time, and a 7.5 cm source-to-sensor distance, comparing otherwise identical acquisitions with and without microlenses.

**Figure 1.**
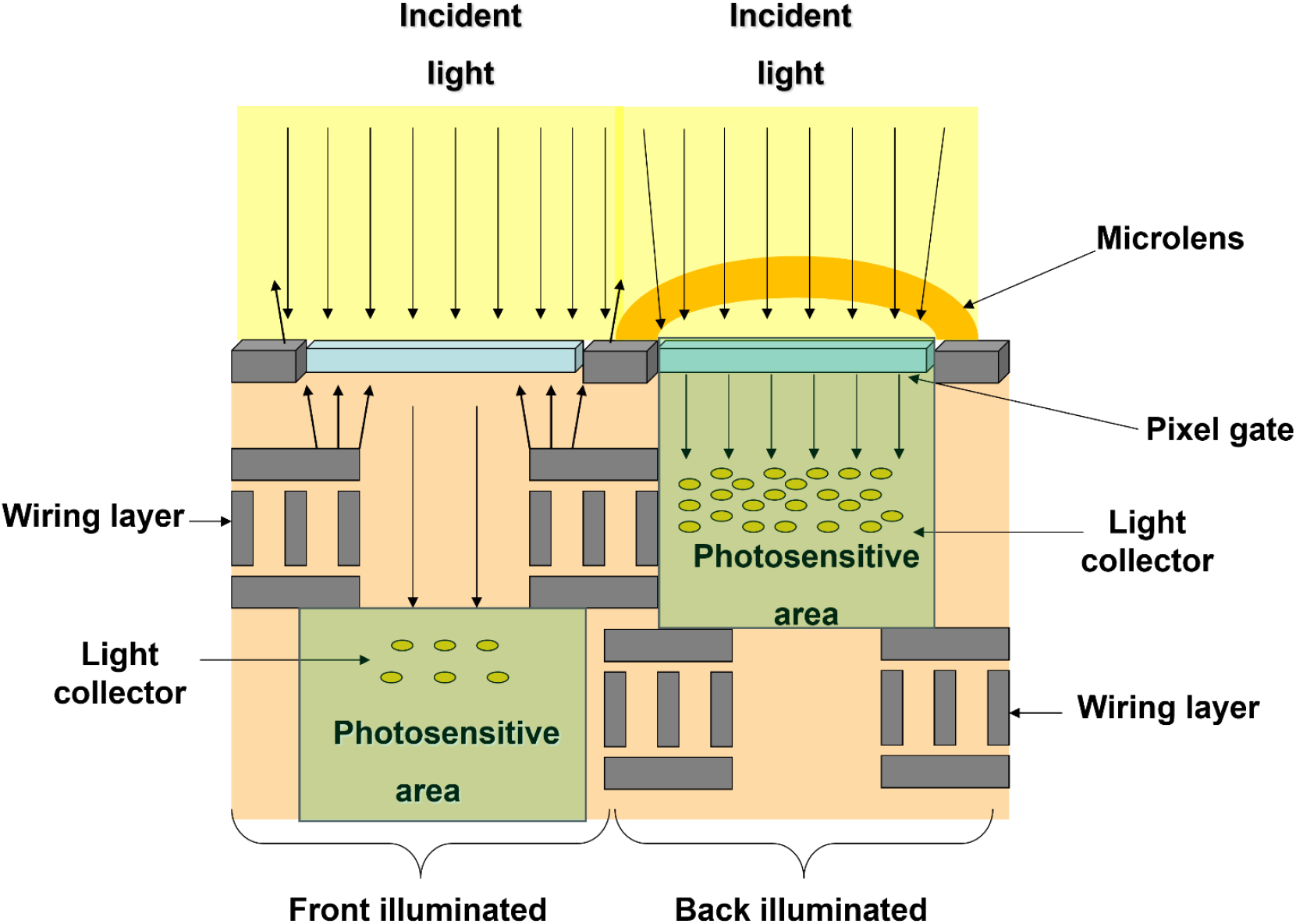
Pixel architecture comparison. Back-illuminated CMOS sensors place the photosensitive area at the entrance surface and use microlenses to guide light into each pixel, increasing quantum efficiency. This BI-M architecture enables 8 µm pixels for high-resolution, low-dose imaging.

**Figure 2.**
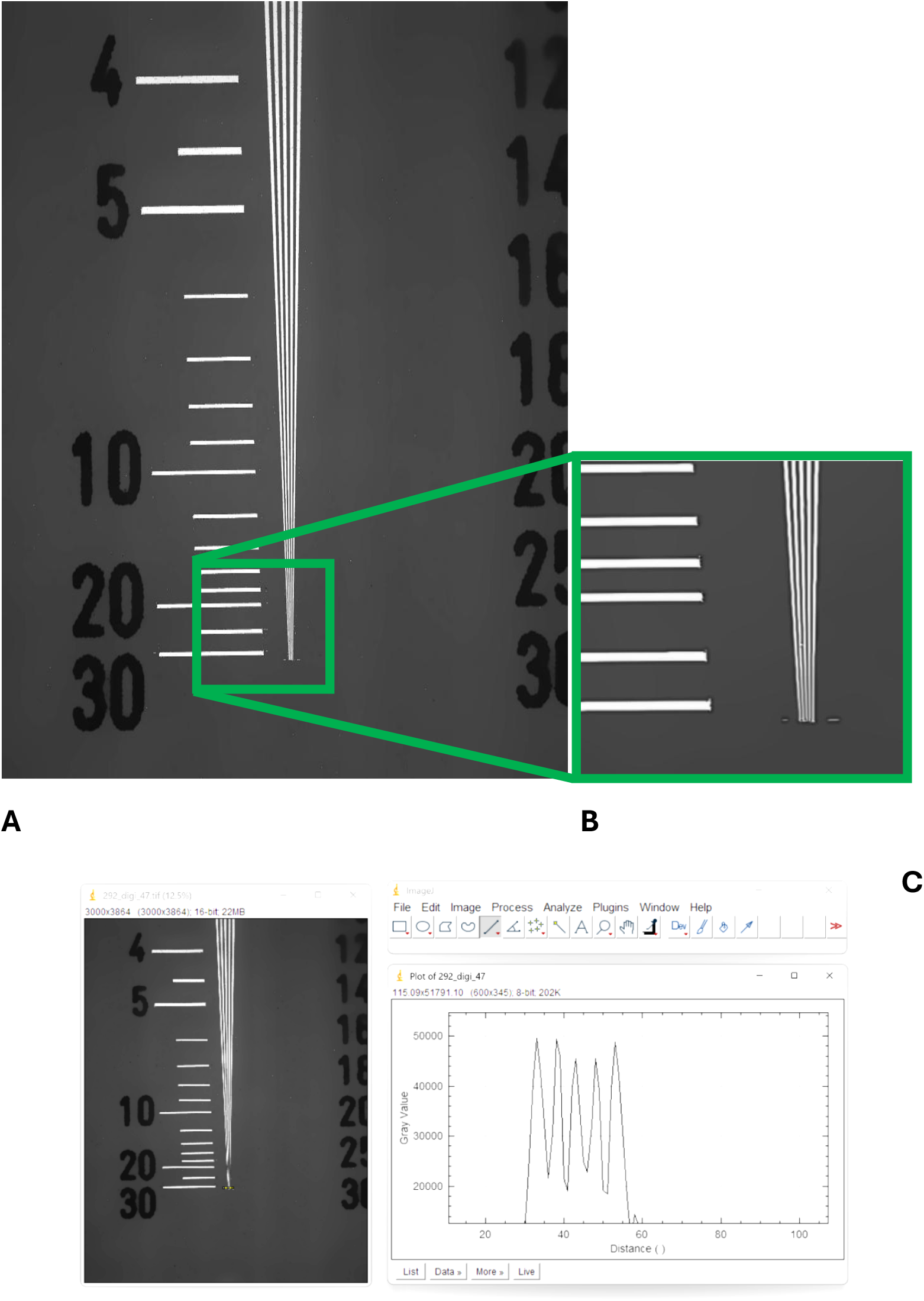
Resolution target. (A) Resolution test pattern acquired with the prototype. (B) Zoomed 30 lp/mm region. (C) Bars corresponding to 30 lp/mm are consistent with ImageJ analysis and the measured MTF results.

## Results

The measured dose rate of 83.6 mGy s^−1^ at 5 cm corresponds to an incident air kerma of 20.9 mGy per 0.25 s exposure and 0.075 mAs. Because the prototype operated at 0.3 mA × 0.25 s = 0.075 mAs, this represents a photon fluence several-fold lower than that used in conventional digital radiography. For comparison, clinical chest and extremity DR exposures typically deliver 0.14– 0.27 mAs per pulse (≈40–60 mGy s^−1^ at 100–120 kVp), while mammography averages 100–120 mAs (≈10–20 mGy per image) at 25–35 kVp, and dental CBCT fields range from 0.1–0.3 mSv (≈10–30 mGy air kerma) depending on field size. The prototype therefore achieved equivalent or superior image resolution at an incident dose 40–70 % lower than DR and orders of magnitude lower tube current (0.3 mA vs 5–300 mA). These findings confirm that the 8 µm BI + M architecture preserves image quality at substantially reduced radiation burden. Extrapolating to 20 cm minimum clinical distances, the dose would scale by the inverse square (≈1/16), yielding an expected ≈1.3 mGy per image, which remains well below routine diagnostic reference levels.^5 8 9 11 12 41 42^

The resolution test pattern (Fig. 2) and the measured MTF (Fig. 3A) showed sustained contrast transfer to 30 lp/mm, more than four times that of radiography panels and three times that of mammography detectors. CT and CBCT performance, constrained to ∼1–2 lp/mm, were significantly lower than 30 lp/mm. This limitation arises from their larger pixel sizes and corresponding Nyquist frequencies, whereas the prototype’s Nyquist frequency of 62.5 lp/mm represented a continuous spatial bandwidth not previously available in clinical detectors. Noise performance is illustrated in Figure 3B. The NPS curve for the prototype extended across the full measured frequency range up to the Nyquist limit. In contrast, clinical reference systems exhibited truncated noise spectra, corresponding to their lower Nyquist frequencies (≈1–6 lp/mm). CT and CBCT also demonstrate relatively greater low-frequency noise, consistent with voxel averaging and coarse reconstruction kernels that emphasize large-scale structure at the expense of fine detail. ^5 7 9 12 41^

**Figure 3.**
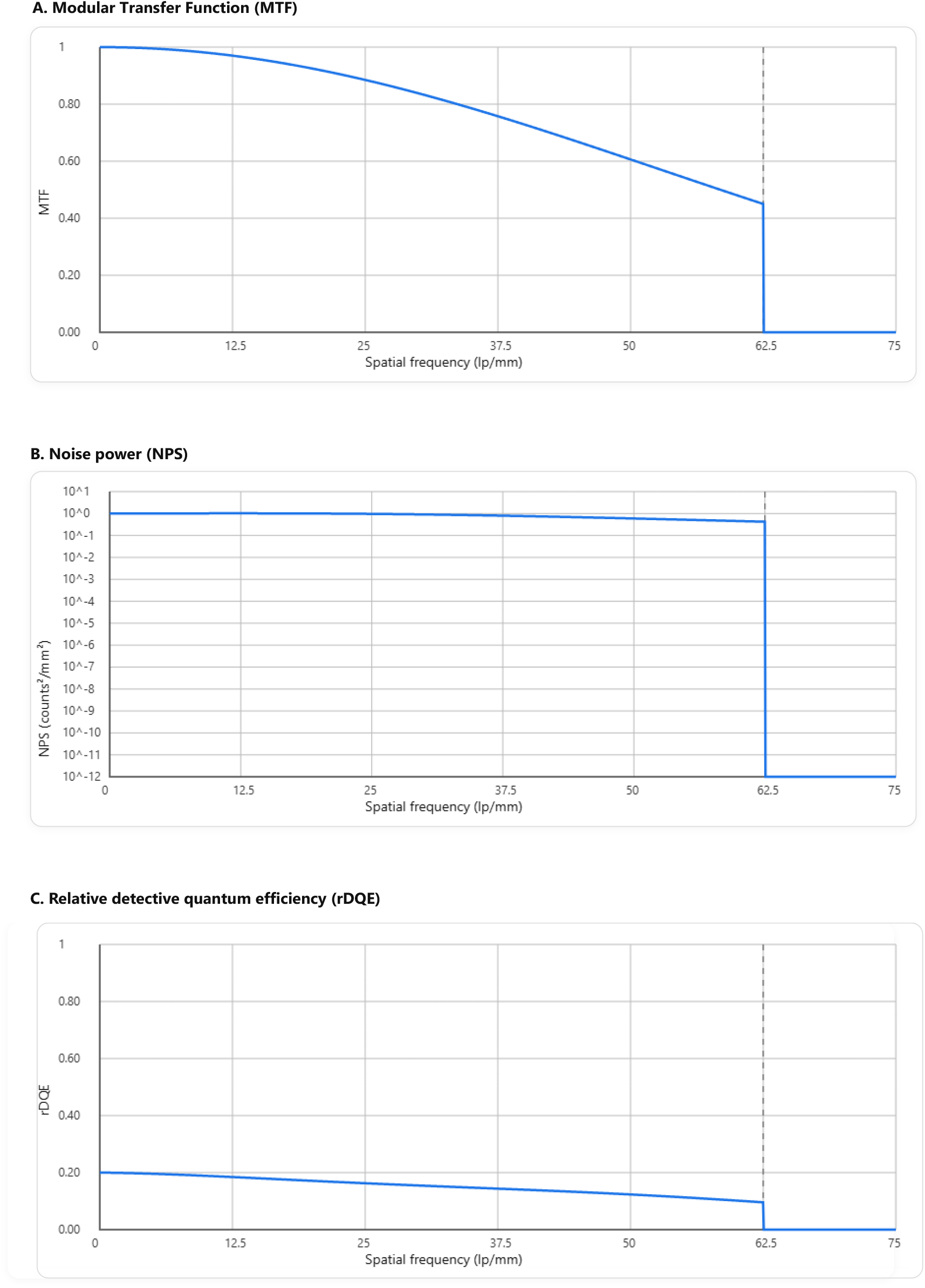
Prototype detector performance. (A) Modulation transfer function (MTF). (B) Noise power spectrum (NPS). (C) Relative detective quantum efficiency (rDQE). Curves are shown up to the prototype Nyquist frequency of 62.5 lp/mm.

Relative DQE results are presented in Figure 3C. The rDQE value remains comparable to the CBCT scan modality. Efficiency remained elevated across the full measured frequency range, preserving performance even at 30 lp/mm. By comparison, radiography/mammography detectors decay to near-zero efficiency by ∼5 lp/mm, and CT/CBCT by ∼1–2 lp/mm. The prototype detector demonstrates a significantly extended MTF compared to DR, mammography, CBCT, and CT comparators, consistent with its smaller pixel size (8 µm, corresponding to a Nyquist frequency of 62.5 lp/mm). However, the prototype rDQE remains below DR [Alikunju 2023], García-Mollá et al. (2011), and Monnin et al. (2020), and comparable to CBCT values [Steiding 2014], but consistent with the lack of optical bonding in this prototype. Future iterations incorporating optical bonding are anticipated to markedly improve rDQE while preserving the high-frequency resolution advantage.^5 11 41^ Relative to the no-microlens configuration, microlenses increased measured MTF by approximately 9% at 15 lp/mm, 28% at 20 lp/mm, 25% at 40 lp/mm, and 16% at 50 lp/mm, corresponding to an overall ∼9% increase in MTF area from 0–50 lp/mm (Figure 4).

**Figure 4.**
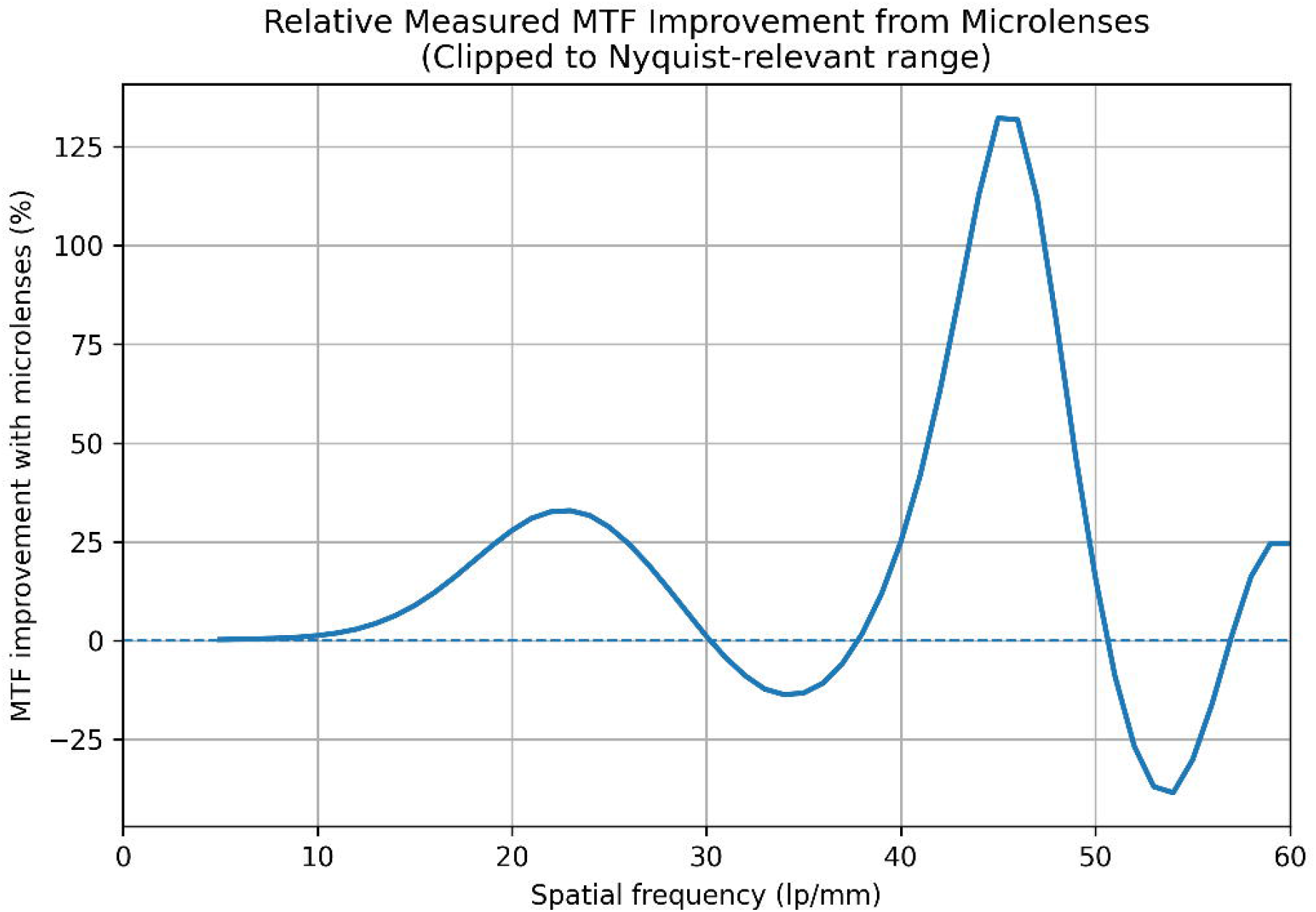
Relative improvement in measured system MTF with microlenses. Relative percentage change in measured system modulation transfer function (MTF) as a function of spatial frequency for an 8 µm back-side illuminated CMOS detector with microlenses compared with an otherwise identical detector without microlenses.

## Discussion

This work represents the first step toward the translation of a novel high-resolution dental CMOS-BI with M detector technology and microfocus x-ray sources into broader medical imaging applications. Results confirmed that the prototype detector overcame long-standing limitations in radiographic imaging. In clinical terms, general DR typically uses 100–180 cm source-to-detector distance, mammography operates at a fixed 65 cm, CT employs a 50 cm source-to-isocenter radius (≈100 cm source-to-detector), and dental or medical CBCT systems range from 15–60 cm, all substantially longer than our 5 cm prototype geometry. When normalized to detector area and patient-equivalent distances, the delivered dose per frame remains an order of magnitude below that used in standard medical modalities. This low-mA regime (≤ 0.3 mA) demonstrates that microfocus sources combined with high-DQE 8 µm pixels can sustain diagnostic image quality without approaching the milliamperes routinely applied in medical systems. Such dose efficiency is central to translating dental microfocus technology to broader medical imaging applications where ALARA compliance and cumulative exposure reduction are critical.^5 8 9 11 12 41 42^

For decades, spatial resolution in X-ray detectors has been dictated by pixel pitch, scintillator blur, and X-ray source focal spot, with little improvement beyond ∼7 lp/mm in clinical practice. These measurements show that by employing 8 µm pixels, a high-resolution LYSO (Ce) scintillator, and a microfocus X-ray source, it is possible to sustain contrast and efficiency well beyond 30 lp/mm. This advance derives not only from pixel size reduction but from the integration of BI pixel architecture and M in combination with a micro focal spot at the source level. Conventional detectors that shrink pixel pitch below 70 µm typically suffer a loss of quantum efficiency, as fewer photons reach the active area. The BI pixel design and M array resolve this limitation by efficiently channeling photons into each 8 µm pixel without striking the wiring layers, preserving sensitivity despite a small pitch. Although the scintillator primarily limits system resolution, microlenses improve the measured system MTF under low-dose imaging conditions by increasing the detected signal and reducing measurement noise. The micro-sized focal spot at the X-ray source (20 μm) reduces the penumbra effect and produces sharp, clear images when compared to the 0.4 mm and larger focal spots used in dental and medical imaging. This synergy enables two benefits: contrast transfer beyond 30 lp/mm and meaningful dose reduction at 0.3 mA. For the prototype image in Figure 2 (0.25 s), we operated at 0.3 mA and 71.7 kVp, yielding 0.075 mAs per image. This is 46% lower than 0.14 mAs and 72% lower than 0.27 mAs, i.e., well below a representative clinical DR per-image range (∼0.14–0.27 mAs) and mammography at 100-120 mAs. On tube current alone, the same 0.3 mA operating point is ≈333–1000× lower than DR, mammography and CT (100–300 mA), and ≈17–67× lower than CBCT (5–20 mA) when beam quality and geometry are held constant. We therefore use mA/mAs strictly in context and focus cross-modality comparisons on image-quality metrics (MTF, NPS, rDQE). Clinically, the MTF results could translate into the ability to visualize micro-calcifications, micro-lesions, fine trabecular structures, and subtle fractures that existing modalities miss. Prototype rDQE stays meaningful up to very high frequencies, while DR/mammography/CBCT/CT loses efficiency early. A high frequency rDQE clinical advantage means the prototype can preserve detail at lower mA or sharper images without increasing dose. The prototype maintained measurable noise power into frequencies far beyond those accessible to conventional systems. That indicates improved noise suppression, which is crucial for low-dose clinical workflows. These results suggest that, with further optimization of detector–scintillator coupling, the prototype could outperform current DR systems in both resolution and efficiency, supporting translation to clinical medical imaging. This detector technology also allows the introduction of microfocus X-ray sources for clinical applications from the industrial and material sciences fields.^40^ Clinically, this combination could reduce oncologic failed detection, particularly for early-stage diagnosis and residual disease detection after therapy, at doses lower than today’s standard practice. In practical terms, it addresses the 56% errors rate in diagnostic radiology, directly mitigating one of the largest vulnerabilities in medicine.^1 2 3 4^ Although the prototype was designed for dental intraoral applications, the technology is inherently scalable. The sensor wafer design allows CMOS design and butting into large flat-panel arrays, providing a pathway to high-resolution detectors for radiography, mammography, extraoral dental, CBCT, and CT.^36^ Such scaling would permit the benefits demonstrated in this prototype to extend across multiple clinical applications, from screening to image-guided interventions.

Because most radiologic cancers are missed at the earliest and most treatable stages, largely due to pixel-size driven resolution limits, an 8 um BI-M architecture capable of sustaining contrast beyond 30 lp/mm could improve visualization of microcalcifications, early tumor margins, and subtle soft-tissue changes. Such capabilities are especially important in breast and lung cancer, where micro-calcifications and micro-nodules are frequently undetected on standard mammogram, radiography and CT/CBCT. By providing high-resolution imaging at substantially reduced mA, this detector architecture may support earlier cancer detection while minimizing cumulative radiation burden.

An initial clinical avenue for translating this dental high-resolution, low-dose detector architecture into medical imaging is pediatric and thin-anatomy radiography. Children present smaller anatomical structures, lower mineralization, and higher radiosensitivity, making them disproportionately affected by pixel size driven resolution limits and dose constraints in current flat-panel detectors. Thin-anatomy regions in adults (e.g., extremities, breast micro-detail, craniofacial imaging) share similar spatial-resolution demands. Because dental detectors are already designed to resolve very fine structures at low doses, this class of applications represents a scientifically and clinically aligned pathway for early adoption of ultra-high-resolution microfocus-based systems. This approach supports improved visualization of subtle findings, reduces cumulative radiation exposure, and aligns with existing dose-reduction campaigns in pediatric imaging.^43 44 45^ These results validate the 8 µm BI with M CMOS detector prototype’s dose efficiency at realistic geometries and indicate that, even when adjusted for standard clinical distances, the projected air-kerma per image remains well below those of conventional digital radiography, mammography, CT, or CBCT systems.

## Conclusion

When adjusted for standard clinical geometries, the projected dose reduction remains significant, confirming that the prototype’s performance translates to realistic imaging distances. The 8 µm BI with M CMOS detector prototype and a microfocus X-ray source achieved 30 lp/mm resolution, continuous low-noise performance across its Nyquist bandwidth, and sustained dose efficiency at high spatial frequencies, compared with MTF, NPS, and DQE measurements of existing medical imaging modalities, including DR, mammography, CT, and CBCT. The prototype delivered superior image quality and efficiency while operating at 0.3 mA. Its BI pixel architecture and M array enabled this performance in combination with a microfocus X-ray source, ensuring both resolution gains and dose reduction. The BI-M architecture broke the dose-resolution trade-off by enabling small pixels without loss of quantum efficiency. Microlenses therefore represent an enabling system-level component for improving measured spatial-frequency response (MTF) in high-resolution, low-dose X-ray imaging architectures. Scalable to larger flat-panel arrays, this technology has the potential to improve early-stage disease and cancer diagnosis, reduce failed detection, and mitigate radiation risk in medical imaging, with pediatric and thin-anatomy imaging serving as the initial clinical translation pathway. More research will be conducted reproducing clinical settings and source-to-object distances used in existing medical modalities via an 8 µm BI with M CMOS large format flat panel detector for microfocus clinical applications.

## Supporting information

30 lp/mm original picture

## Data Availability

All data produced in this study are available from the authors upon reasonable request.

