## Supplementary figures and images for "From Dental to Medical Imaging: Translational 8 μm Pixel Size, Low-dose and Ultra-High-Definition X-ray Detector for Microfocus Clinical Applications"

### 30 lp/mm original picture

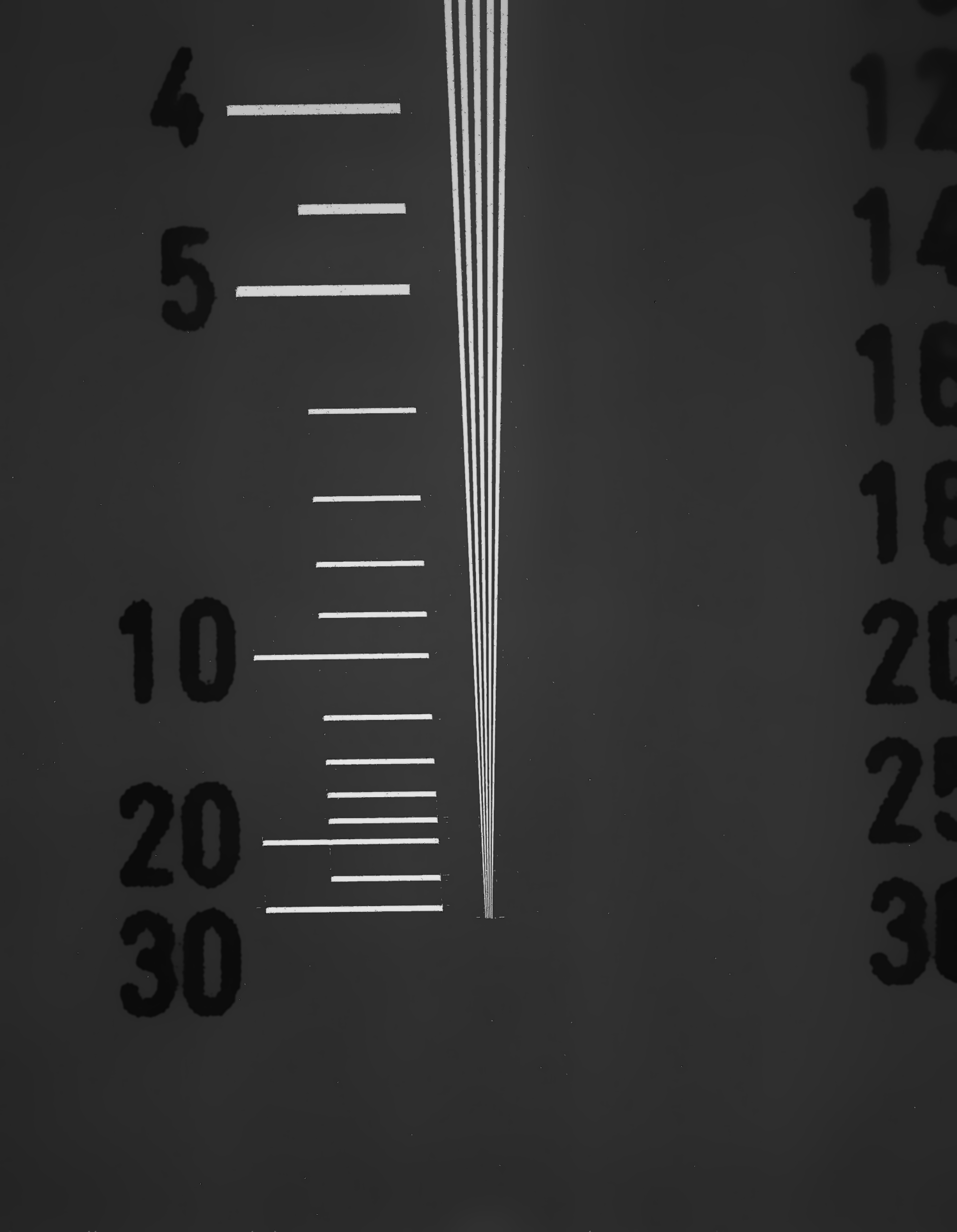
